# An Extended Laboratory Validation Study and Comparative Performance Evaluation of the Abbott ID NOW™ COVID-19 Assay in a Coastal California Tertiary Care Medical Center

**DOI:** 10.1101/2020.06.14.20130518

**Authors:** Stewart Comer, David Fisk

**Affiliations:** Santa Barbara Cottage Hospital, Laboratory Department, Santa Barbara, CA, USA; Santa Barbara Cottage Hospital, Infection Control Physician, Santa Barbara, CA, USA

**Author notes:** **Corresponding Author:** Stewart Comer MD FCAP.

## Abstract

The Abbott ID NOW™ COVID-19 assay is a rapid molecular diagnostic test particularly designed for on-site, rapid turnaround point of care (POC) testing. The utilization of rapid diagnostic tests is integral to optimizing workflow within the hospital and/or procedural-based clinics. The capability to provide both rapid disposition and correct patient classification during this COVID-19 pandemic is critically important with broad infection control implications for both patients and healthcare staff. A tightly controlled, extended laboratory validation was performed at our medical center to determine the negative test agreement of the Abbott ID NOW™ compared with the BD MAX™ analyzer, a laboratory-based, two target, molecular analyzer with a sensitive cycle threshold (Ct) positive cutoff value of ≤ 42. There was strict adoption of the procedures listed in the Abbott ID NOW™ Instruction for Use (IFU)^1^ insert delineating preferred practices for “optimal test performance.” Under these conditions, our institution demonstrated a significant negative percent agreement with 116 out of 117 patients correlating, which equates to a 99.1% concordance similar to a recently reported correlation study2.

## Introduction

The Abbott ID NOW™ COVID-19 assay, which received Food and Drug Administration Emergency Use Authorization (FDA EUA) approval on 27 March 2020, is a rapid molecular diagnostic test particularly designed for on-site, rapid turnaround point of care (POC) testing. The analyzer uses isothermal nucleic acid amplification of a single unique region of the RNA-dependent RNA polymerase (RdRp) genome using a self-contained, combined collection and testing kit. Although the system can utilize a variety of throat, nasal or nasopharyngeal swabs with specific allowance for storage stability, the Instruction for Use (IFU) ^1^ states that “optimal test performance” is achieved utilizing the swabs provided with the collection kit and “should be tested as soon as possible after collection.” By mid-April 2020, Cottage Health (CH) adopted the use of the Abbott ID NOW™ analyzer and validated utilizing archived positive and negative specimens in viral transport media (VTM) as well as direct smears from known positive and negative patients. Our validation with direct smears demonstrated 100% concordance; however, there was only 80% concordance with the archived VTM specimens. Our analysis of this false negative discordance was coincident with the published studies from Northwell Health^3^ and media reports about Cleveland Clinic^4^ highlighting this issue. Given the proposed use of the Abbott ID NOW™ to test predominantly symptomatic hospital admissions in the context of the uncertainty imparted by these studies, Cottage Health purposely designed a process using dry swabs only with immediate on-site testing to comprehensively ascertain the actual negative test performance of the Abbott ID NOW™.

## Study Design and Methods

The key conceptual elements of this prospective validation study were originally designed by CH senior infectious disease physicians and operationalized by emergency department (ED) and laboratory leadership. This study was initiated with the general design to test all COVID-19 symptomatic prospective hospital admissions in the ED with the Abbott ID NOW™ and, if negative, recollect expeditiously and test on a laboratory-based molecular analyzer. The medical center in which this study was performed is a major referral center for Santa Barbara and adjoining counties in mid-coastal California. At the inception of this extended validation study, the county had just reached its peak and the laboratory-tabulated prevalence of COVID-19 was slightly above 10%, which according to the Infectious Disease Society of America (IDSA) guidelines would have been classified as high prevalence. As the peak subsided the final average over the time interval of the study demonstrated an intermediate prevalence5 of 5.2%. As such, the Abbott ID NOW™ with their proprietary nasal swabs was the frontline testing platform and the Becton Dickinson BD MAX™ selected as the laboratory-based platform. To optimize clinical test sensitivity, particularly in the absence of published data on comparative swab sensitivity for COVID-19 detection, the decision was made to adopt the practice of collecting combined nasopharyngeal (NP) and oropharyngeal (OP) swabs. Another important design feature is that the Abbott ID NOW™ was collected and tested immediately on-site in the ED diminishing any significant testing delay and reducing any potential RNA degradation associated with room-temperature, dry swab storage. Finally, a cardinal design feature to this prospective study was the deliberate recollection to occur within a short time interval (measured in single digit hours) in order to focus specifically on testing the negative agreement concordance between the two platforms rather than test clinical sensitivity variability associated with swabs for common respiratory viruses^6^. This is addressed in accordance with the IDSA guidelines by repeat testing 24 – 48 hours after an initial negative test on patients with intermediate to high clinical suspicion for COVID-19. The latter issue of enhancing clinical sensitivity for SARS-Cov-2 virus is addressed by repeat testing after 24 hours; whereas, our approach was intended to remove potentially confounding results from the types of retrospective studies that compare the Abbott ID NOW™ on initial testing with subsequent laboratory-based molecular analyzers greater than 24 hours after admission. This comparison can be flawed since it effectively removes important design controls that could be the source of discordance associated with temporal heterogeneity inherent with the SARS-Cov-2 virus. Our study was designed to specifically focus on the comparative performance of the two different testing analyzers.

## Results

From 16 April 2020 to 21 May 2020, a total of 117 patients were collected and the patients ranged from 11 to 97 years, representing 56% male, 44% female and having a mean age of 61 years. The dual swab collection for the second test was obtained within an average of 2.2 hours with 40% collected in less than 1 hour from the original Abbott ID NOW™ collection. There was a significant negative percent agreement with 116 out of 117 patients correlating for a 99.1% concordance.

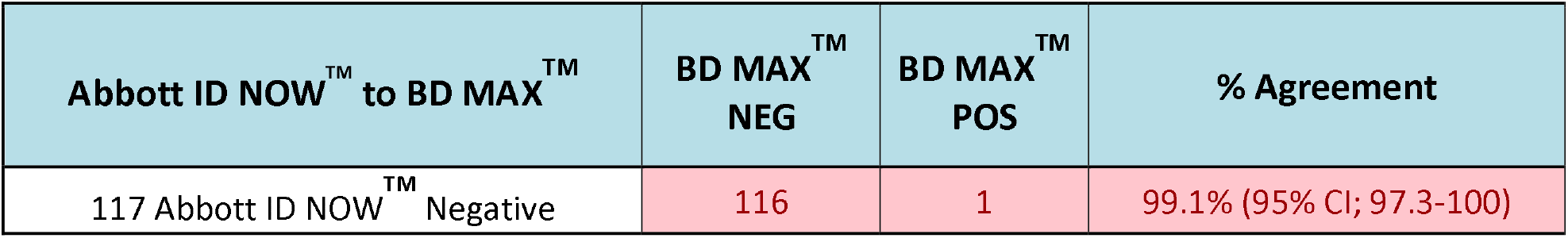

## Discussion

The intent of this comparative validation study has been to construct a simple design of experiment including strict adherence to the Abbott ID NOW™ procedures delineated in the IFU1 as being optimal and exclude preanalytic confounding variables of specimen stability and swab type. This was accomplished by the following practices to optimize test sensitivity, specifically: 1) use of the “nasal swab accompanying the test kit” and not a substitute, 2) transport the swab dry to prevent any dilutional effect, 3) test the swab immediately on site and not introduce delays to minimize in vitro RNA degradation, and 4) optimize swab transfer to the receiver reservoir without creating air bubbles and verify optimal fluid dispensing to the transfer cartridge. Our results demonstrate a significant correlation of the Abbott ID NOW™, a single target, rapid diagnostic POC analyzer with the BD MAX™ analyzer, a laboratory-based, two target, molecular analyzer with a sensitive cycle threshold (Ct) positive cutoff value of ≤ 42. The use of combined NP and OP specimens was adopted in this extended validation study to enhance preanalytic clinical sensitivity. The cohort of 117 patients entering the tertiary care medical center was predominantly symptomatic but also included some asymptomatic patients although this is likely irrelevant since the COVID-19 IDSA guideline panel quote many studies that claim essential equivalency of test accuracy for symptomatic and asymptomatic patient populations^5^.

## Conclusion

Our institutional results are correlated with a recently reported correlation study^2^ performed from 8-22 April 2020 at the Everett Clinic, Washington in which the Abbott ID NOW™ was compared to the Hologic Panther Fusion®, a laboratory-based, two target, molecular analyzer. The concordance rate of negative results was determined to be 99.8% with 932 out of 934 Abbott ID NOW ^M^ negative results concurring with the comparator platform, relatively similar to our concordance rate of 99.1%. This Everett Clinic study, which consisted of a much larger sample size, has correspondingly more robust statistical power in comparison to our study. Our comparative false negative agreement stands in contrast to much lower concordance rates with a slightly smaller sample reported by another recent institutional study^7^ using the dry nasal swab, but were stored at room temperatures for 1 to 2 hours prior to testing. A possible conclusion to our institutional laboratory validation is that additional studies should be directed toward further investigations comparing variable room temperature stability time intervals with particular attention focused on the collection to actual test performance time interval on the Abbott ID NOW™ compared to an FDA EUA-approved, laboratory-based analyzer.

## Data Availability

All data will be available. There is no supplemental data being submitted.

## Acknowledgements

The following clinical leaders from Cottage Health Infectious Disease, Emergency Medicine and Laboratory Departments provided invaluable insights and support that operationalized this prospective, extended laboratory validation study. Lynn Fitzgibbons MD (Infectious Disease, Santa Barbara Cottage Hospital); Brett Wilson MD and Bridget Crooks RN, MSN (Emergency Medicine Department, Santa Barbara Cottage Hospital); Lynette Hansen PhD, CLS, MT (ASCP), Charlene Fernandez CLS, MT (ASCP) and Jane Choe MBA, CLS, MT (ASCP) (Laboratory Department, Santa Barbara Cottage Hospital and Pacific Diagnostic Laboratories).

## Notes

**Conflict of Interest Statement:** No conflicts of interest.

**Funding/Sponsor:** There is no funding beyond the normal and customary costs associated with performing required laboratory validation studies, which is supported by the performing laboratory test (Cottage Health - Pacific Diagnostics Laboratories).

### Competing Interest Statement

The authors have declared no competing interest.

### Clinical Trial

None

### Funding Statement

Funding/Sponsor: There is no funding beyond the normal and customary costs associated with performing required laboratory validation studies, which is supported by the performing laboratory test (Cottage Health - Pacific Diagnostics Laboratories).

### Author Declarations

Approved by Cottage Health IRB on 4 JUN 2020 with IRB #20-80mx.

